# The impact of the COVID-19 pandemic on mental health in the general population – a comparison between Germany and the UK

**DOI:** 10.1101/2020.08.27.20182980

**Authors:** Franziska Knolle, Lisa Ronan, Graham K. Murray

## Abstract

**Background:** The COVID-19 pandemic has led to dramatic social and economic changes in daily life. First studies report an impact on mental health of the general population showing increased levels of anxiety, stress and depression. In this study, we compared the impact of the pandemic on two culturally and economically similar European countries: the UK and Germany.

**Methods:** Participants (UK=241, German=541) completed an online-survey assessing COVID-19 exposure, impact on financial situation and work, substance and media consumption, mental health using the tSymptom-Check-List-27 (SCL-27) and the Schizotypal Personality Questionnaire.

**Results:** We found distinct differences between the two countries. UK responders reported a stronger direct impact on health, financial situation and families. UK responders had higher clinical scores on the SCL-27, and higher prevalence. Interestingly, German responders were less hopeful for an end of the pandemic and more concerned about their life-stability.

**Conclusion:** As 25% of both German and UK responders reported a subjective worsening of the general psychological symptoms and 20-50% of German and UK responders reached the clinical cut-off for depressive and dysthymic symptoms as well as anxieties, it specifically shows the need for tailored intervention systems to support large proportions of the general public.

## Introduction

The world health organisation (WHO) declared the outbreak of the coronavirus disease 2019 SARS-CoV-2 (COVID-19) a pandemic on March 11, 2020 (WHO-Media-Briefing). In order to slow down a rapid spread across and within countries, many government responded with strict measures, including lockdown with school and work-place closures, self-isolation and social distancing, border closures, restrictions of travel, to reduce the transmission of the virus. On March 18, 2020 the WHO published a statement presenting mental health and psychosocial considerations for the general public, acknowledging the potential impact of this public health emergency on mental health of the general population [2]. As the COVID-19 outbreak compared to other recent pandemics or medical emergencies is much larger in scale, its consequences are unpreceded and therefore more difficult to predict. The seriousness of the measures taken to control the outbreak have led to immediate and serious concerns on mental health of the general society [3] with calls for urgent and direct actions [4]. From former epidemics, as recently reviewed by Brooks and colleagues [5], such as the 2003 epidemic of the severe acute respiratory syndrome (SARS) or the 2014 outbreak of Ebola, we know that quarantine, isolation and social distancing is related to anxiety, depression, sleep disorders, etc. In the current pandemic, however, entire countries were locked-down for much longer periods of time. Increase of job insecurity and economic hardship [6, 7], as well as domestic violence [8, 9], substance abuse [10] and media consumption [11, 12] have been discussed as risk factors for impacting mental health. First studies (e.g. [13–15]) confirm increased levels of stress, anxiety, depressive symptoms, sleep disorders as well as an increase in suicidal ideation, etc, also in the current pandemic.

In response to the outbreak and spread of the pandemic, different countries even within Europe followed different strategies. Germany went into lockdown rapidly and managed to control the increase of infections effectively, whereas the UK due to a delayed lockdown faced a much higher plateau (see Figure 1)[16] which also led to an increase in numbers of deaths that were at the end of April 2020 20% higher than predicted, whereas Germany was nearly 3% lower than expected. Balmford and colleagues (2020) used epidemiological models to estimate the “price of life” that various nations were willing to pay in order to protect their people. According to their estimation the German government was prepared to pay a factor of 10 more per life than the UK, in April 2020. It is to be expected that these different strategies and governmental choices have an impact on the nation’s mental well-being. We were therefore interesting in comparing the mental health impact of the pandemic on the general population of the UK and Germany, using an online survey investigating the impact on life circumstances and assessing mental health with two different psychological questionnaires (Symptom Check List, SCL-27; Schizotypal Personality Questionnaire, SPQ). We hypothesised that responders of both nations, UK and Germany, would report an increase in psychological symptoms, but that the increase would be stronger in UK responders. We supplemented the general mental health questionnaire SCL-27 with the SPQ, as we reasoned that a potential increase in anxiety and distress could be accompanied by an increase in psychotic-like experiences [17] that could be captured in the SPQ.

**Figure 1.**
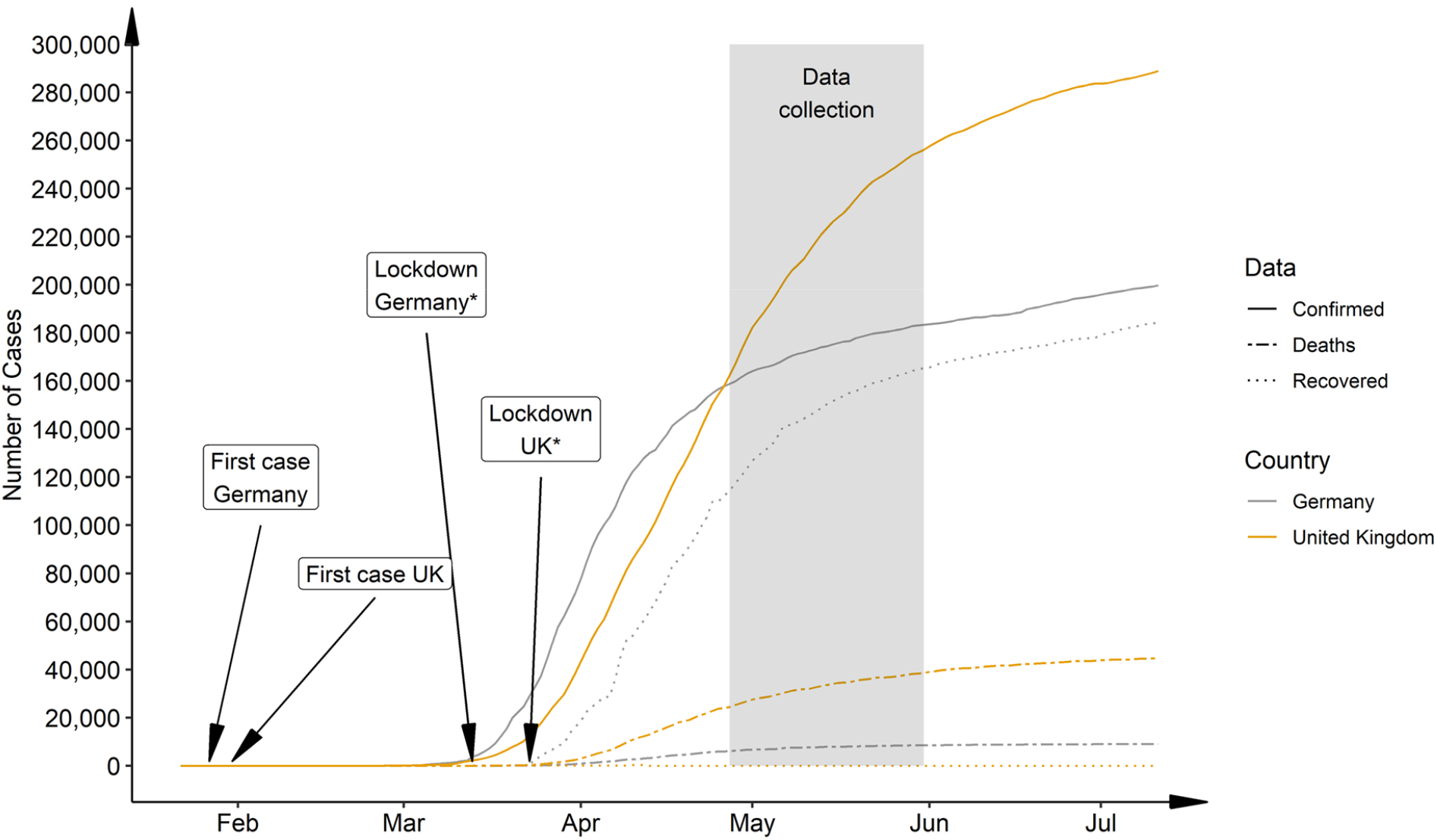
National progression of COVID-19 cases, deaths and recoveries comparing Germany and the UK from Jan. 22,2020 – Jul. 11,2020. Recovery rate UK: after April 12., 2020 recovered cases are not reported for the UK. *Germany followed a state-wise lockdown, with the first state going in lock-down on Mar.13,2020 and the last state on the Mar. 16, 2020. UK announced nationwide lockdown on Mar.23, 2020. Data taken from the 2019 Novel CoronaVirus CoViD-19 (2019-nCoV) Data Repository by Johns Hopkins University Center for Systems Science and Engineering (JHU CSSE) (https://github.com/CSSEGISandData/COVID-19) on Jul. 11, 2020

## Methods

### Study design and procedure

The questionnaire assessing mental and physical health and COVID-19 exposure was designed as an online survey using EvaSys (https://www.evasys.de, Electric Paper Evaluationssysteme GmbH, Luneburg, Germany). The questionnaire was available in German and English. For participant recruitment we used a snowball sampling strategy to reach the general public. Data collection took place from 27/04/2020-31/05/2020. The completion of the survey took approximately 35 min. Participation was voluntary. Participants did not receive any compensation.

Ethical approval was obtained from the Ethical Commission Board of the Technical University Munich (250/20 S). All participants provided informed consent.

### Outcome

The survey consisted of three parts. The first part, partially comprised of the Coronavirus Health Impact Survey (CRISIS, http://www.crisissurvey.org/), which assessed demographics, COVID-19 exposure (infection status, symptoms, contact), mental and physical health questions. In the second part, we assessed the general mental health status (global severity of symptom index (GSI-27)) using the Symptom Check List (SCL) with 27 items [18, 19] and its subdimensions. For all SCL-items we recorded the subjective change during the pandemic compared to before. In the third part, using the Schizotypy Personality Questionnaire (SPQ, [20]) we evaluated total schizotypic symptoms (SPQ-total), subdimensions [21], and subjective change per item.

### Statistical analysis

Statistical analysis and visualisations were computed using R (RStudio Team (2020). RStudio: Integrated Development for R. RStudio, PBC, Boston, MA URL http://www.rstudio.com/.). We first describe demographics and COVID-19 exposure variables, using non-parametric analysis. For the country comparison we used a chi-square test or a Wilcox test for categorical and continuous variables respectively to explore differences between the groups on the demographics and the COVID-19 exposure variables.

To further explore the differences between the UK and Germany and timepoints in the CRISIS variables, we conducted robust ANOVAs [22] with country (UK, Germany) and timepoint (before pandemic (i.e., subjective rating) and during pandemic) as between-subjects factor.

To identify possible predictors for worse or better functioning, we furthermore applied multivariate Poisson regression models to assess the associations between the outcome and the predictor variables. Our outcome variables were continuous scores measured using the SCL-27 and the SPQ. We investigated total score (SPQ-total, GSI-27) as well as subscales for the SPQ and the SCL-27.

## Results

### Demographics

The survey was complete by 860 participants. Two participants did not provide consent and were excluded. 6 participants did not consent to sharing the data publicly, and will be removed from the open-access data set. In this paper we focus on the comparison of respondents resident in the UK (N=239) and in Germany (N=541). All descriptive and statistical results are described in Table 1.

**Table 1.** Cohort demographics and COVID-19 exposure including impact on life; and differences between Germany and the

|  |  | Whole sample | UK | Germany | Chi² / Wilcox for country comparison: UK vs. GER |
| --- | --- | --- | --- | --- | --- |
| N |  | 858 | 239 | 541 | X-squared =118.2, df = 1, p-value < 0.001 *** |
| Age |  | 43.27yrs (SD15.4) | 39.01yrs (SD16.0) | 45.36yrs (SD14.8) | Wilcox: 48386 p-value < 0.001 *** |
| Gender | Female | 71.6% | 73.6% | 71.2% | X-squared = 0.36, df = 2, p-value = 0.8 |
|  | Male | 25.4% | 24.3% | 25.9% |  |
|  | Diverse | 0.5% | 0.4% | 0.6% |  |
|  | Missing | 2.5% | 1.7% | 2.4% |  |
| Education | School leavers | 0.1% | 0.4% | - | X-squared = 69.3, df = 7, p-value<0.001*** |
|  | 8years – A-Levels | 14.3% | 19.2% | 13.1% |  |
|  | Professional college or Bachelor | 24.3% | 31.8% | 21.6% |  |
|  | Masters or higher | 60.1% | 47.3% | 64.9% |  |
|  | Missing | 1.2% | 1.3% | 0.4% |  |
| Children at home (max.18years) | Yes | 28.1% | 21.1% | 30.7% | X-squared = 7.83, df = 1, p-value = 0.005 ** |
|  | Missing | 2.0% | 1.6% | 2.6% |  |
| COVID19 exposure |  |  |  |  |  |
| Suspected infection | Positive Test | 0.1% | - | 0.2% | X-squared = 7.25, df = 3, p-value = 0.06 |
|  | Diagnosis | 1.3% | 2.5% | 0.7% |  |
|  | Symptoms | 15.6% | 18.8% | 14.2% |  |
|  | No infection | 82.2% | 78.7% | 83.9% |  |
|  | Missing | 0.8% | - | 0.9% |  |
| Symptoms | Fever | 8.5% | 10.0% | 8.0% | X-squared = 1.04, df = 1, p-value = 0.3 (based on ‘no symptoms’) |
|  | Cough | 17.1% | 21.3% | 15.5% |  |
|  | Shortness of breath | 9.4% | 10.0% | 9.8% |  |
|  | Sore throat | 21.2% | 23.4% | 19.9% |  |
|  | Fatigue | 27.4% | 28.5% | 27.7% |  |
|  | Lost smell/taste | 2.9% | 4.2% | 2.2% |  |
|  | Infected eyes | 3.7% | 3.8% | 3.5% |  |
|  | Other symptoms | 6.3% | 6.3% | 6.7% |  |
|  | No symptoms | 55.2% | 51.9% | 55.8% |  |
| Contact to people with potential infection | Positive test | 7.8% | 3.3% | 9.8% | X-squared = 1.14, df = 1, p-value = 0.3 (based on ‘no contact’) |
|  | Diagnosis | 2.1% | 5.4% | 0.6% |  |
|  | Symptoms | 12.8% | 17.2% | 11.7% |  |
|  | No contact | 78.1% | 74.9% | 78.4% |  |
| Impact on work and financial situation |  |  |  |  |  |
|  | Home office | 48.7% | 50.6% | 46.8% | X-squared = 2.39, |
| Impact on work | Reductions of hours | 7.1% | 6.7% | 7.4% | df = 1,<br>p-value = 0.1<br>(based on 'no change') |
|  | Unpaid leave | 2.6% | 3.4% | 2.4% |  |
|  | Overtime/negative hours<sup>‡</sup> | 7.7% | 1.3% | 11.3% |  |
|  | Lost job | 3.9% | 2.9% | 3.7% |  |
|  | No change | 16.1% | 13.0% | 17.4% |  |
| Impact on family | Infected | 5.8 | 10.5% | 4.4% | X-squared = 38.3,<br>df = 1,<br>p-value < 0.001 ***<br>(based on 'no impact') |
|  | Hospitalised | 1.8% | 1.7% | 2.0% |  |
|  | Death | 1.3% | 3.8% | 0.4% |  |
|  | Quarantine, symptoms | 15.3% | 23.9% | 12.2% |  |
|  | Quarantine, no symptoms | 7.5% | 14.6% | 4.6% |  |
|  | Reduced working hours | 16.7% | 26.4% | 12.6% |  |
|  | Lost job | 6.8% | 10.9% | 4.4% |  |
|  | No impact | 65.9% | 49.0% | 71.9% |  |
| Financial impact of COVID-19 | No impact | 54.0% | 46.0% | 58.8% | X-squared = 19.2,<br>df = 4,<br>p-value <0.001 *** |
|  | Slight | 16.4% | 23.4% | 12.8% |  |
|  | Moderate | 13.3% | 11.7% | 12.8% |  |
|  | Big | 12.6% | 15.9% | 11.7% |  |
|  | Extreme | 3.3% | 2.9% | 3.3% |  |
|  | Missing | 0.5% | - | 0.7% |  |
| Not enough money for food | Yes | 2.9% | 2.9% | 3.1% | X-squared = 0.03,<br>df = 1,<br>p-value = 0.8 |
|  | Missing | 1.1% | 1.0% | 1.0% |  |
| Personal judgement of the situation |  |  |  |  |  |
| Are the restrictions stressful? | Not at all | 13.5% | 11.3% | 14.8% | X-squared = 6.3,<br>df = 4,<br>p-value = 0.2 |
|  | Slightly | 26.8% | 29.3% | 26.3% |  |
|  | Moderately | 27.6% | 25.5% | 28.1% |  |
|  | Very | 20.5% | 24.3% | 18.7% |  |
|  | Extremely | 11.3% | 9.2% | 12.2% |  |
|  | Missing | 0.2% | 0.4% | - |  |
| How concerned are you about your life stability? | Not at all | 34.6% | 46.4% | 30.5% | X-squared = 21.7,df = 4, p-value < 0.001*** |
|  | Slightly | 23.4% | 22.2% | 23.8% |  |
|  | Moderately | 19.1% | 14.2% | 21.3% |  |
|  | Very | 15.4% | 10.9% | 17.2% |  |
|  | Extremely | 6.8% | 5.0% | 6.8% |  |
|  | Missing | 0.7% | 1.3% | 0.4% |  |
| How hopeful are you of a soon end of the pandemic in your region of residence? | Not at all | 17.1% | 14.2% | 17.9% | X-squared = 28.824,<br>df = 4, p-value = 8.489e-06 *** |
|  | Slightly | 32.1% | 27.6% | 34.9% |  |
|  | Moderately | 28.6% | 25.2% | 30.1% |  |
|  | Very | 14.7% | 19.7% | 12.8% |  |
|  | Extremely | 7.6% | 13.0% | 4.3% |  |
| Does the pandemic have any | No | 35.0% | 31.0% | 36.4% | X-squared = 14.225,<br>df = 2, p-value = 0.0008147*** |
|  | Few | 35.6% | 29.3% | 37.7% |  |
|  | Some | 29.1% | 38.9% | 25.9% |  |
| positive impact on your personal life? | Missing | 0.4% | 0.8% | - |  |
| Mental and physical health status |  |  |  |  |  |
| Self-rated mental health, before COVID-19 | Excellent | 15.0% | 14.2% | 15.3% | X-squared = 34.938, df = 4, p-value = 4.783e-07 *** |
|  | Very good | 32.4% | 21.8% | 37.0% |  |
|  | Good | 28.7% | 30.1% | 28.3% |  |
|  | Fair | 16.4% | 21.3% | 14.6% |  |
|  | Poor | 5.8% | 11.3% | 3.3% |  |
|  | Missing | 1.6% | 1.3% | 1.5% |  |
| Regular treatment for mental illness, before pandemic | Yes | 13.3% | 17.6% | 10.1% | X-squared = 8.7436, df = 1, p-value = 0.003107 ** |
|  | Missing | 11.4% | 13.4% | 7.2% |  |
| Continuation of treatment during pandemic | More | 2.3% | 3.6% | 1.0% | X-squared = 5.3799, df = 2, p-value = 0.06789 |
|  | Less | 12.5% | 13.6% | 11.5% |  |
|  | Same | 85.3% | 82.8% | 87.8% |  |
| Self-rated physical health, before pandemic | Excellent | 12.1% | 13.0% | 11.3% | X-squared = 1.5322, df = 4, p-value = 0.8209 |
|  | Very good | 33.1% | 32.2% | 33.3% |  |
|  | Good | 33.9% | 31.8% | 35.7% |  |
|  | Fair | 16.3% | 17.2% | 15.5% |  |
|  | Poor | 3.2% | 3.8% | 3.3% |  |
|  | Missing | 1.4% | 2.1% | 1.0% |  |
| Regular treatment for physical illness, before pandemic | Yes | 19.5% | 18.4% | 20.2% | X-squared = 0.0008792, df = 1, p-value = 0.9763 |
|  | Missing | 11.6% | 14.6% | 7.0% |  |
| Continuation of treatment during pandemic | More | 1.0% | 1.8% | - | X-squared = 5.7522, df = 2, p-value = 0.05635 |
|  | Less | 14.1% | 14.3% | 14.4% |  |
|  | Same | 85.0% | 83.9% | 85.6% |  |
| *$\leq .05$, **$\leq .01$, ***$\leq .001$; *Overtime/negative hours , is a concept uncommon in the UK. | | | | | |

### COVID-19 Exposure, Impact and Personal Judgement

To explore differences between the countries we used chi-square tests. All results on COVID-19 Exposure, specifically on infection rates (Suppl.Fig.3), symptoms (Suppl.Fig4), contacts (Suppl.Fig.5), work impact (Suppl.Fig.6), family impact (Suppl.Fig.7), and financial impact are presented in Table 1, COVID-19 Exposure.

In general the restrictions were rated similarly stressful between the two countries. UK responders were less concerned about their overall life stability. UK responders were also more hopeful that the pandemic in their region would soon be under control. All findings are presented in Table 1, Personal judgement of the situation.

UK responders rated their mental health status lower compared to the German responders. Also more UK responders received regular treatment for their mental illnesses before the pandemic. The treatment was continued similarly during the pandemic across the two countries. Physical health was judged similarly across the two countries. Regular treatment for physical illnesses was similar between the two countries and the treatment continued in a similar fashion. All findings are presented in Table 1, Mental and physical health status.

*Self-report on sleep, mental health, exercise/outdoor activities, media consumption, and substance/alcohol consumption before and during the pandemic*

Investigating the differences between the countries and timepoints with regard to the CRISIS variables, we applied robust ANOVAs. Means, main effects and interactions for all variables are presented in Table 2.

**Table 2.**
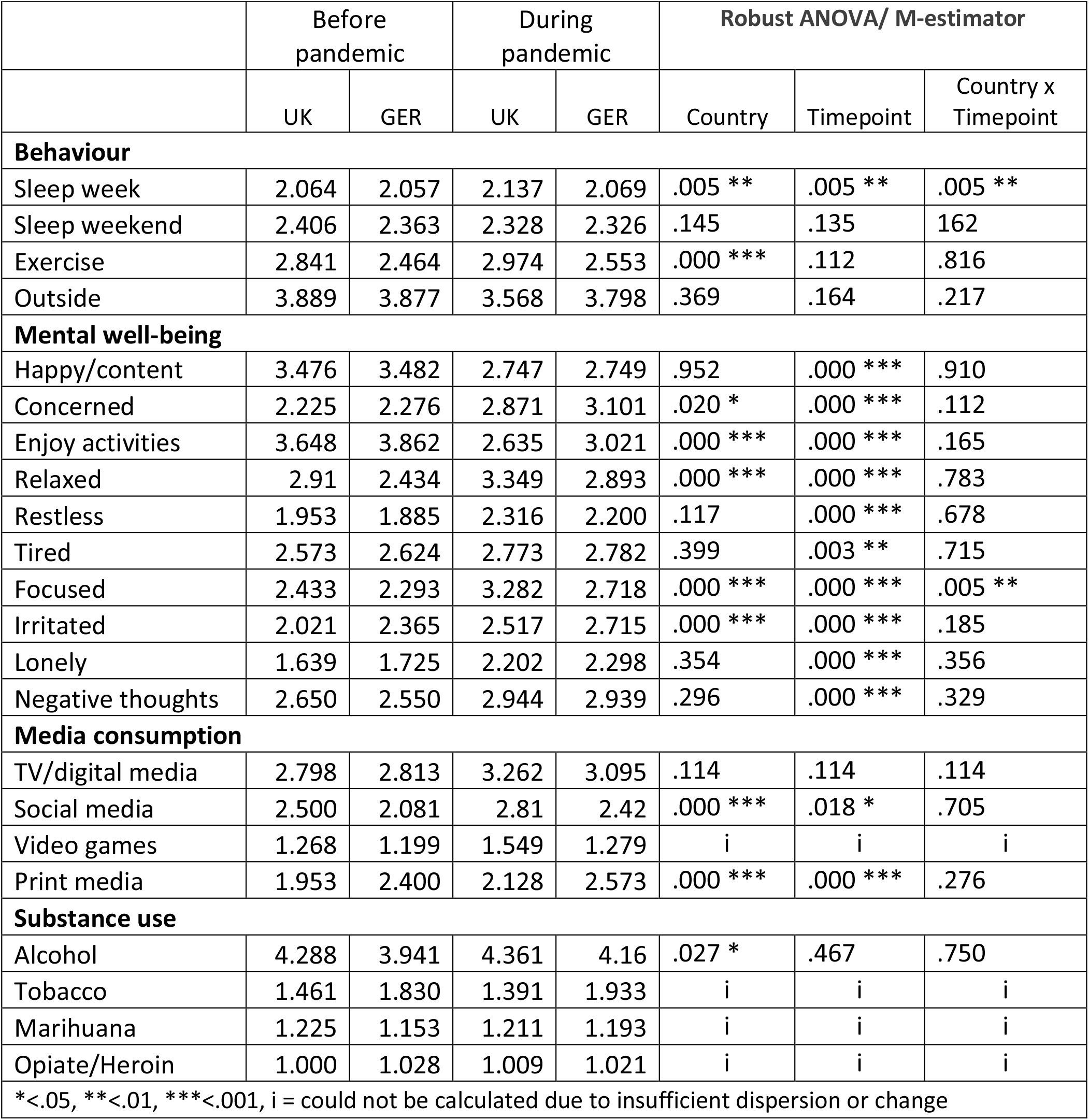
Results from Robust ANOVAs showing the effects of differences between countries (UK and Germany) and time points (before and during the pandemic) on a set of different variables.

|  | Before pandemic |  | During pandemic |  | Robust ANOVA/ M-estimator |  |  |
| --- | --- | --- | --- | --- | --- | --- | --- |
|  | UK | GER | UK | GER | Country | Timepoint | Country x Timepoint |
| **Behaviour** |  |  |  |  |  |  |  |
| Sleep week | 2.064 | 2.057 | 2.137 | 2.069 | .005 ** | .005 ** | .005 ** |
| Sleep weekend | 2.406 | 2.363 | 2.328 | 2.326 | .145 | .135 | .162 |
| Exercise | 2.841 | 2.464 | 2.974 | 2.553 | .000 *** | .112 | .816 |
| Outside | 3.889 | 3.877 | 3.568 | 3.798 | .369 | .164 | .217 |
| **Mental well-being** |  |  |  |  |  |  |  |
| Happy/content | 3.476 | 3.482 | 2.747 | 2.749 | .952 | .000 *** | .910 |
| Concerned | 2.225 | 2.276 | 2.871 | 3.101 | .020 * | .000 *** | .112 |
| Enjoy activities | 3.648 | 3.862 | 2.635 | 3.021 | .000 *** | .000 *** | .165 |
| Relaxed | 2.91 | 2.434 | 3.349 | 2.893 | .000 *** | .000 *** | .783 |
| Restless | 1.953 | 1.885 | 2.316 | 2.200 | .117 | .000 *** | .678 |
| Tired | 2.573 | 2.624 | 2.773 | 2.782 | .399 | .003 ** | .715 |
| Focused | 2.433 | 2.293 | 3.282 | 2.718 | .000 *** | .000 *** | .005 ** |
| Irritated | 2.021 | 2.365 | 2.517 | 2.715 | .000 *** | .000 *** | .185 |
| Lonely | 1.639 | 1.725 | 2.202 | 2.298 | .354 | .000 *** | .356 |
| Negative thoughts | 2.650 | 2.550 | 2.944 | 2.939 | .296 | .000 *** | .329 |
| **Media consumption** |  |  |  |  |  |  |  |
| TV/digital media | 2.798 | 2.813 | 3.262 | 3.095 | .114 | .114 | .114 |
| Social media | 2.500 | 2.081 | 2.81 | 2.42 | .000 *** | .018 * | .705 |
| Video games | 1.268 | 1.199 | 1.549 | 1.279 | i | i | i |
| Print media | 1.953 | 2.400 | 2.128 | 2.573 | .000 *** | .000 *** | .276 |
| **Substance use** |  |  |  |  |  |  |  |
| Alcohol | 4.288 | 3.941 | 4.361 | 4.16 | .027 * | .467 | .750 |
| Tobacco | 1.461 | 1.830 | 1.391 | 1.933 | i | i | i |
| Marihuana | 1.225 | 1.153 | 1.211 | 1.193 | i | i | i |
| Opiate/Heroin | 1.000 | 1.028 | 1.009 | 1.021 | i | i | i |
| * < .05, ** < .01, *** < .001, i = could not be calculated due to insufficient dispersion or change |  |  |  |  |  |  |  |

For the variable *sleep*, sleeping patterns differ during the week differed significantly between the countries (p=0.005) and the timepoints (p=0.005), and in an interaction effect of country by time point (p=0.005).

The amount of *exercise* differed significantly between the countries (p<0.001), but there is no significant change during the pandemic or an interaction.

Analysing *media consumption*, for use of social media and print media we found significant country (both p<0.001) and time-point (p=0.18 and p<0.001, respectively) effects. We did not find differences in the consumption of TV and digital media. Differences for consumption of video games could not be calculated due to insufficient endorsement.

For the *consumption of substances*, consumption patterns of *alcohol* differed significantly different between countries (p=0.027) but no time-point effect or interaction was found. Differences for consumption of *tobacco, marihuana* and *heroin/opiates* could not be calculated due to insufficient endorsement or change.

Regarding self-reported *mental well-being*, there was an effect of time point (min. p=0.003) for all items and a country effect for *concerned* (p=0.02), *enjoy activities* (p<0.001), *relaxed* (p<0.001), *focused* (p<0.001), and *irritated* (p<0.001). For *focused*, scores changed differently between the two countries from before to during the pandemic (p=0.005).

*Effects of the pandemic on psychological symptoms using the Symptom Check List – 27-Point (SCL-27)*

### General Symptom Index

A Wilcox-test for non-parametric data, showed that the distribution of the GSI was different between the two countries (W=84062, p-value = 1.515e-11, 95%-confidence interval: 0.148-0.296; sample estimate for location difference: 0.222). See Figure 2A.

**Figure 2.**
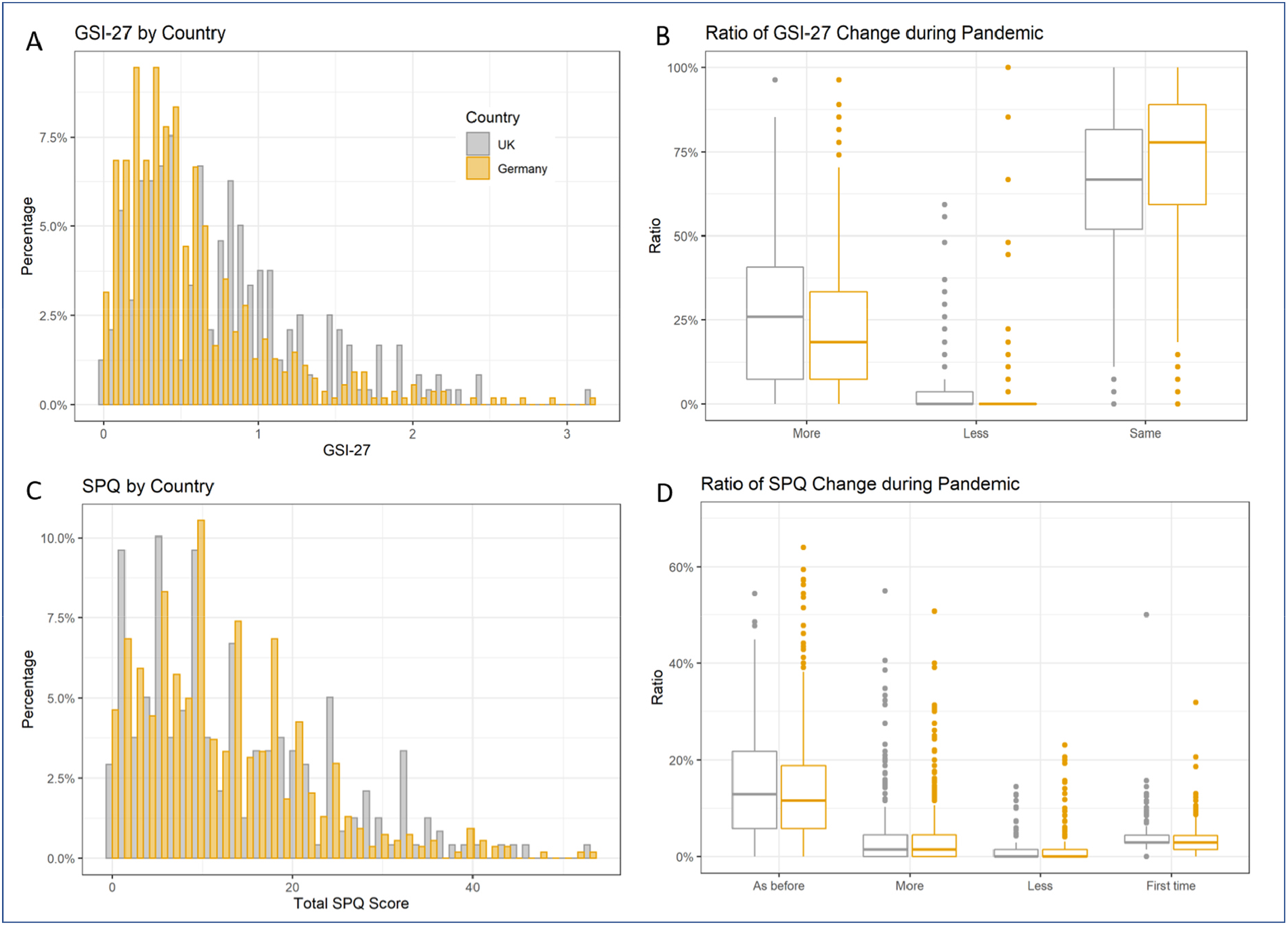
Display of clinical mental health scores measured with the SCL-27 and the SPQ. A: Histogram of distribution of the global severity index based on 27 items (GSI-27) for psychological symptoms, separately shown for countries. B: Boxplot shows the subjective change of global symptom index during the pandemic measured with the SCL, separately for Germany and the UK. C: Histogram of distribution of the total schizotypal personality score (SPQ_total), separately shown for countries. D: Boxplot shows the subjective change of schizotypy symptoms during the pandemic measured with the SPQ-scale, separately for Germany and the UK.

### Clinical cut-off for SLC sub-dimensions and subjective change

In the general population [23], 10-15% of the screened population reach the clinical cut-off on the different sub-dimensions, and require additional investigation. For the sub-dimension of dysthymic symptoms (DYS) 68.5% of the UK responders and 37.6% of the German responders lay above the clinical cut-off; for depressive symptoms it was 48.7% for the UK and 33.5% for the German responders; for symptoms of social phobia 37.1% for the UK and 24.9% for German responders; for symptoms of mistrust 28.9% for the UK and 26.6% for the German sample; for agoraphobic symptoms 43.5% UK and 19.3% for the German responders; and for the vegetative symptoms 19.8% for the UK and 9.6% for the German responders (see Suppl.Fig.8).

We furthermore recorded a subjective rating of change by asking responders on each question of the SCL-27, whether or not this feeling has stay the same, has increased or decreased during the pandemic. In the UK sample 27.0% of the responders reported an increase of symptoms, 3.6% a decrease and 64.1% reported that symptoms stayed the same; whereas in the German sample 22.8% reported more symptoms, 2.5% less and 71.7% the same amount of symptoms (Figure 2B).

*Effects of the pandemic on the schizotypal personality traits using the Schizotypal Personality Questionnaire (SPQ)*

### Total SPQ and subject change during the pandemic

Using a Wilcox-test for non-parametric data, we found that the distribution of the total SPQ-score was not different between the two countries (W=68110, p-value = 0.23). See Figure 2C.

Also for the SPQ, we recorded a subjective rating of change by asking responders on each question, whether or not this feeling/situation has stay the same, has increased or decreased, or has occurred the first time during the pandemic. In the UK sample, 14.7% of the responders reported symptoms as before, 4.8% reported an increase, 1.2% a decrease and 4.4% an occurrence for the first time; similarly, in the German sample, 14.2% reported that symptoms stayed the same, 4.1% reported an increase, 1% a decrease and 3.5% an occurrence for the first time. See Figure 2D. The subjective change of all sub-dimensions is presented in Suppl.Fig.9.

### Association between demographic variables, variables of substance use, media use, sleep, and clinical scores

In order to investigate predictive factors among demographic variables, variables describing exercise, sleep, etc, contributing to clinical scores we conducted two sets of logarithmically normalised multivariate Poisson regression analysis – one set using the GSI and the SCL-subdimensions as outcome variables, and the second set using total SPQ score and the subdimensions as outcome variables. All associations are described in Table 3A and Table4A. In summary for the GSI, we found that responders from Germany have a significantly lower GSI score; female responders are more strongly affected, as well as people with higher consumption rates of marihuana; people who use more social media. Interestingly, people who sleep more (>8h) during week nights have lower GSI scores as well as people who spend more time outside. The predictors and risk factors shift slightly depending on the different subdimensions, but the overall picture is similar.

**Table 3.**
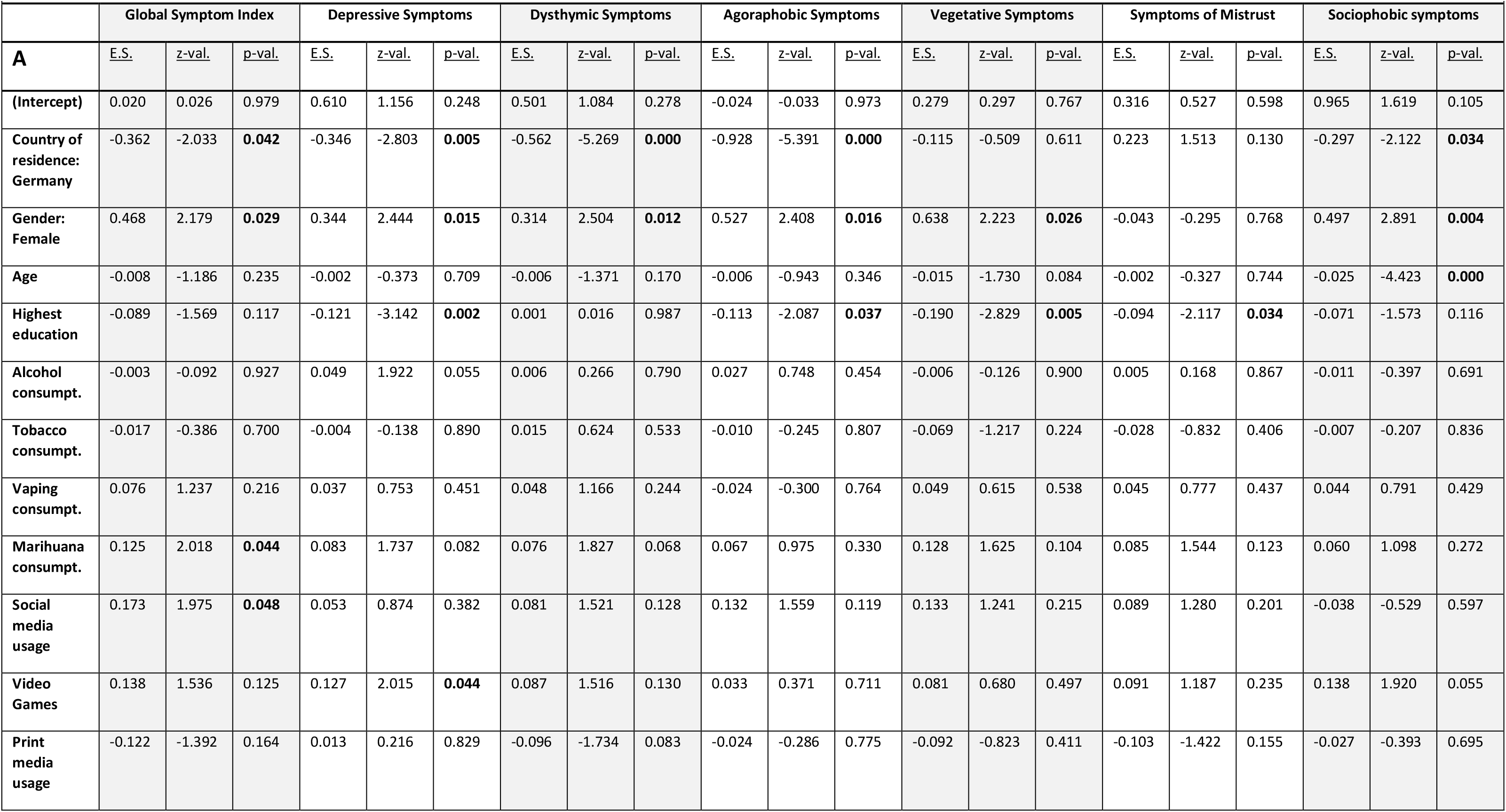
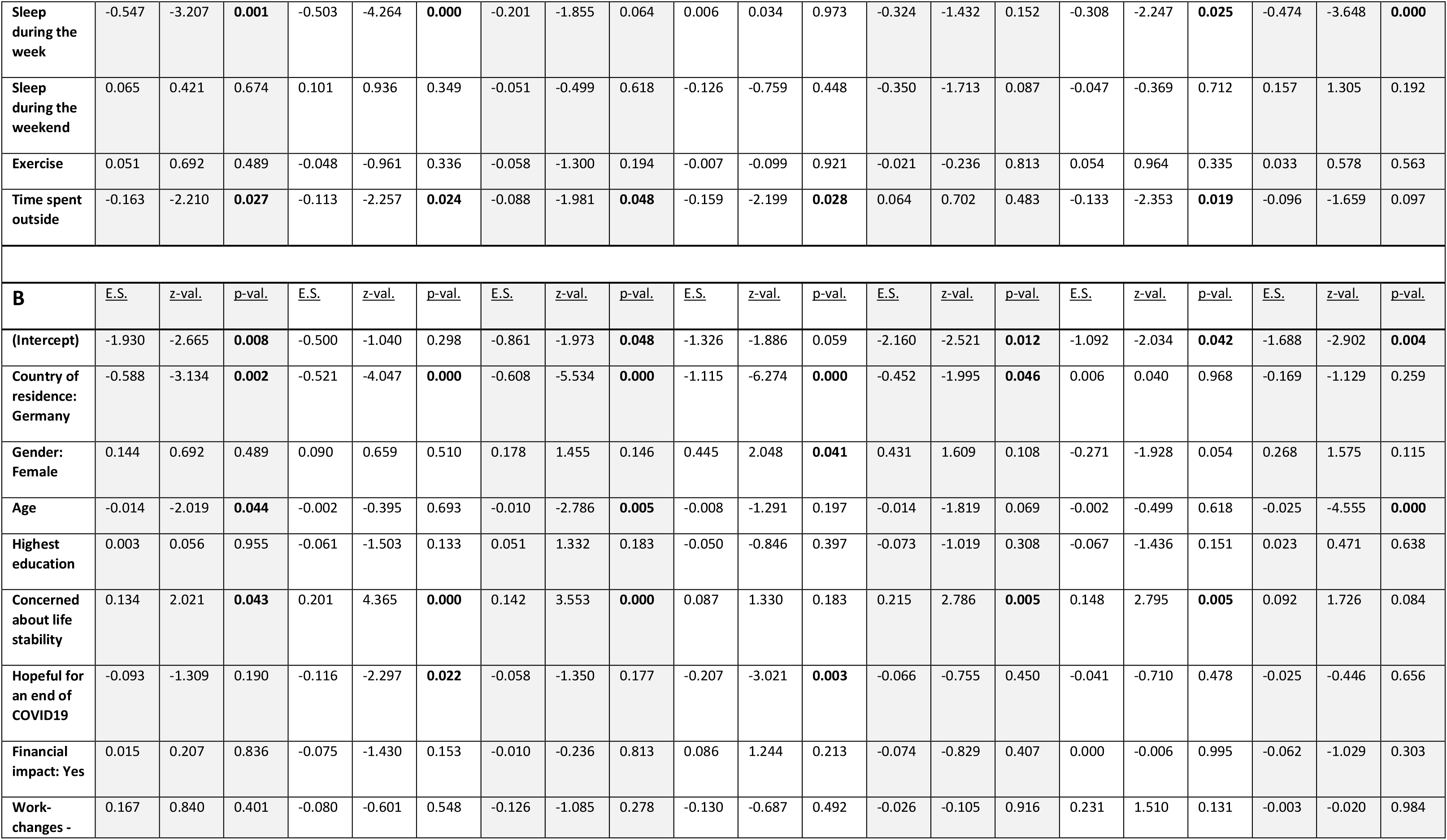
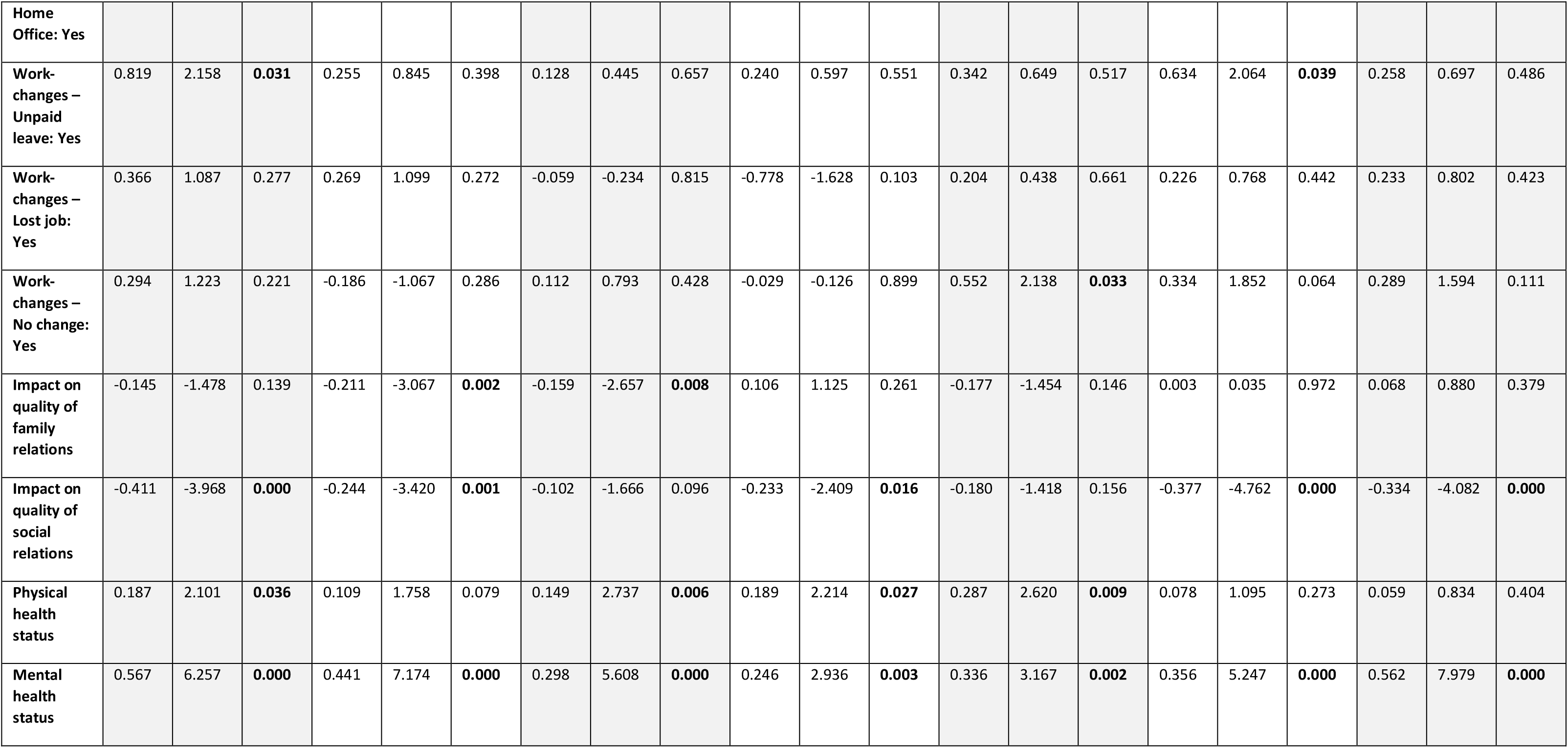
Results for two different logistic regression models, investigating the association between demographic variables, variables of substance use, media use and sleep (A) as well as COVID19 impact (B) and the psychological symptoms (SCL dimensions) as well as global mental health status (GSI).

Interestingly, for the total SPQ score, there was no association with country or gender, instead we saw a protective association with being older, as well as with having a better education. Apart from alcohol consumption, which has a protective association, increased use of tobacco, vaping and marihuana was associated with a higher SPQ scores, and so was the use of social media and video games. The increased use of print media however was associated with lower SPQ scores. Similarly to the associations with GSI, more sleep during the week and the weekend and more time spent outside had a protective association with the total SPQ.

### Association between COVID-19 impact measures, judgement as well as health, and clinical scores

Again we conducted two sets of logarithmically normalised multivariate Poisson regression analyses – one set using the GSI and the SCL-subdimensions as outcome variables, and the second set using total SPQ score and the subdimensions as outcome variables. All associations are described in Table 3B and Table4B respectively. People who were more concerned about their life stability showed a higher GSI. Unsurprising, but with a very strong effect, people who report poor mental health prior to the pandemic were more strongly affected; interestingly the same is true for people who reported low physical health. A protective association is seen for people whose quality of social relationships had not been affected much by the pandemic. Again a similar picture with some variations becomes apparent for the subdimensions. See Table3B.

For the total SPQ score, in this analysis, as opposed to the previous statistical model, female gender was associated with a higher risk. Whereas increased hopefulness for the pandemic to end in the near future was protective, the concern about life stability was a risk factor. People who have been more strongly financially impacted showed higher SPQ-total scores. As also seen for the GSI, people who reported poor mental health prior to the pandemic were more strongly affected; the same is true for people who reported low physical health. Interestingly, whereas working in a home office or being on unpaid leave has a protective effect on the SPQ-total, people who did not see any change in their workplace were also associated with higher total SPQ scores. See Table 4B.

**Table 4.**
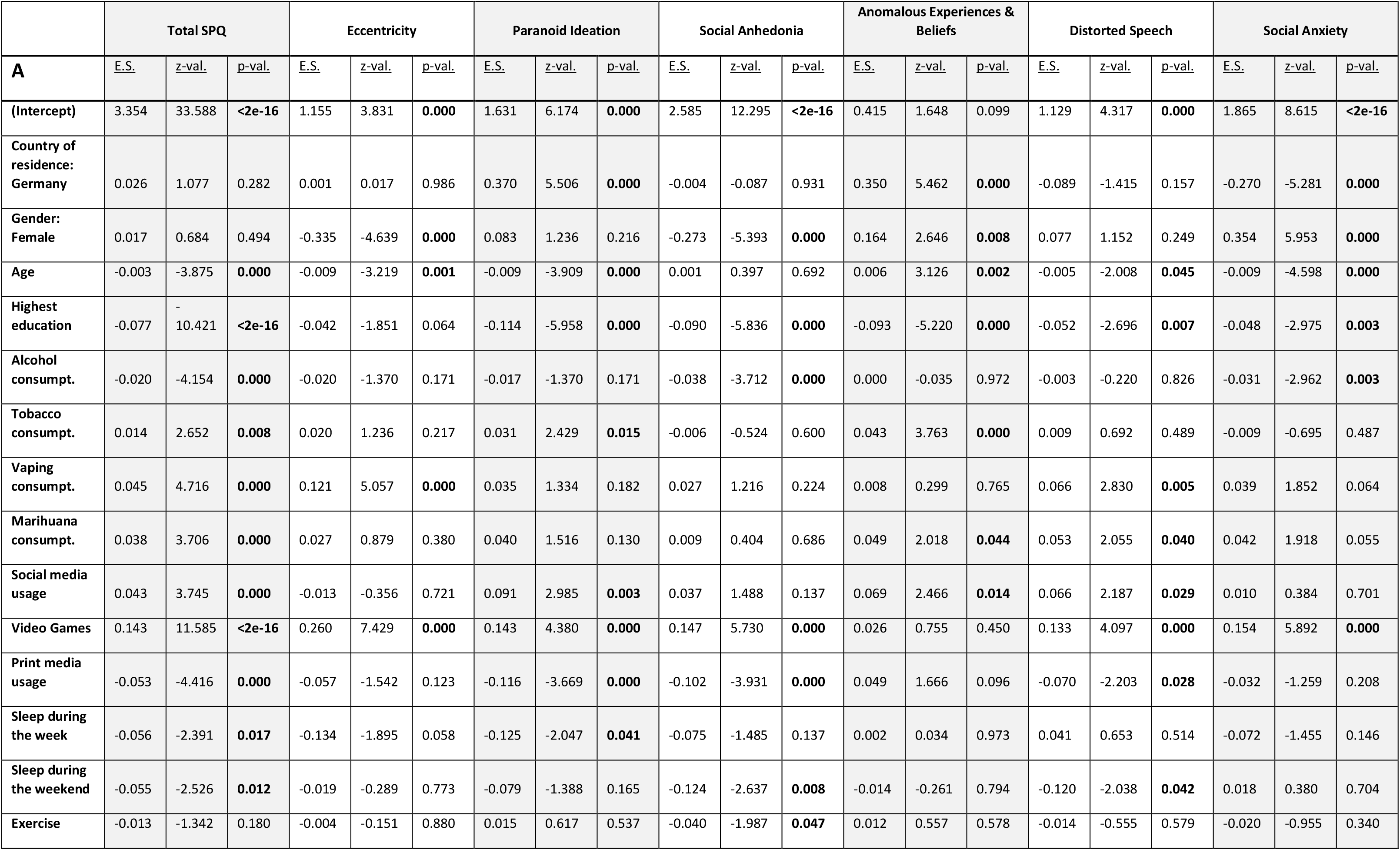
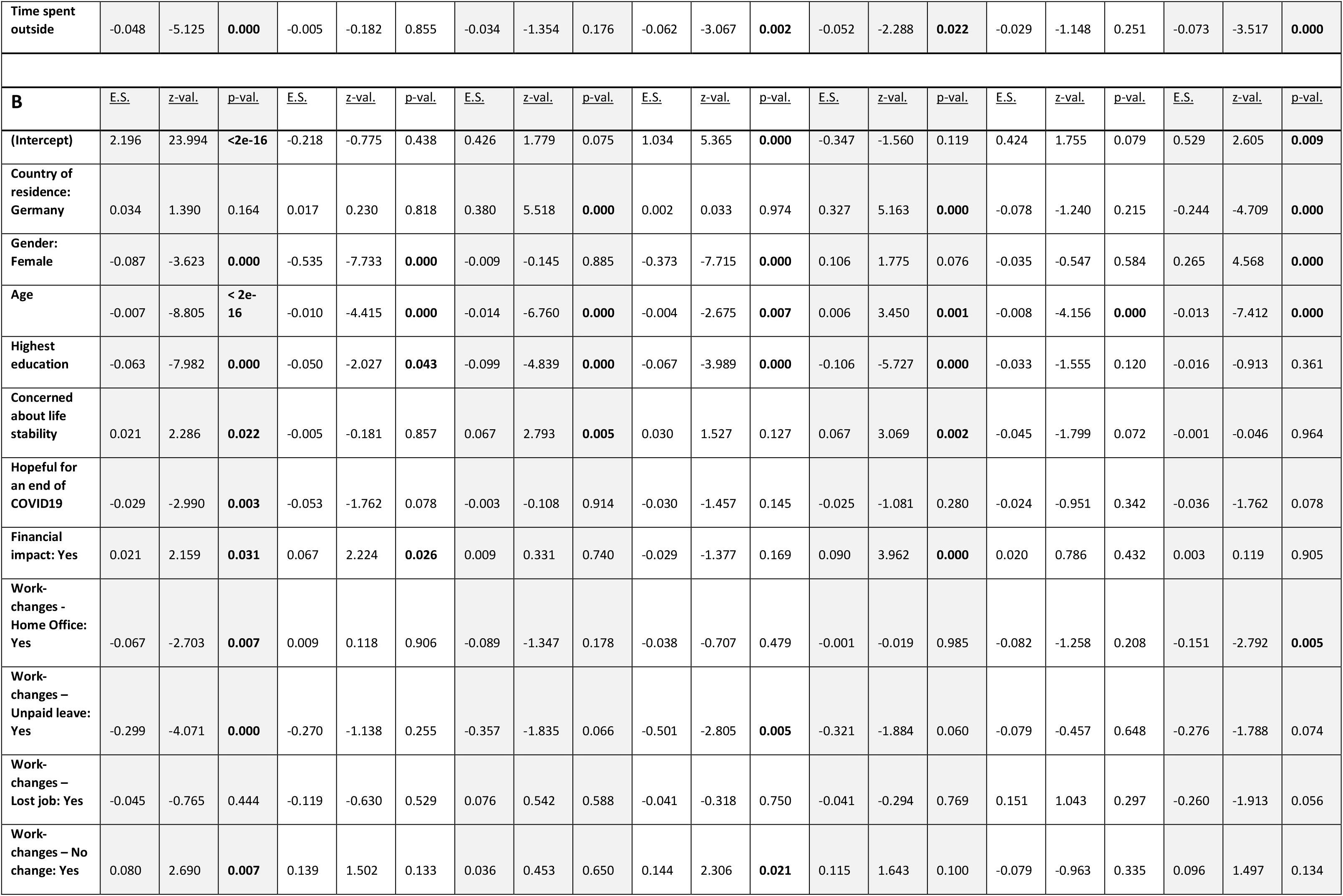
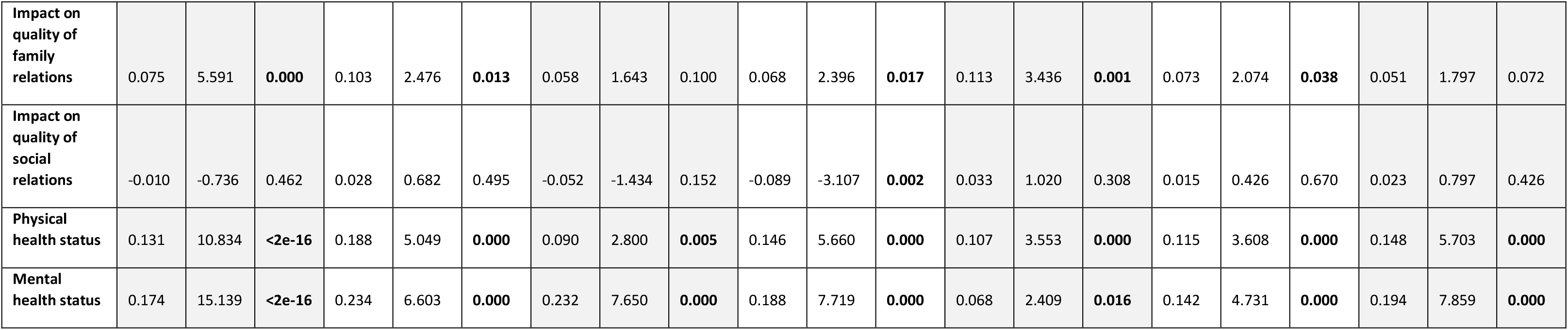
Results for two different logistic regression models, investigating the association between demographic variables, variables of substance use, media use and sleep (A) as well as COVID19 impact (B) and the total Schizotypal Personality Score (Total SPQ) as well as its dimensions.

## Discussion

This study investigated the difference between the impact of the COVID-19 pandemic on the UK and Germany. The impact was assessed using an online survey including questions on the impact on life circumstances, as well as two psychological questionnaires, the Symptom Check List (SCL-27) and the Schizotypal Personality Questionnaire (SPQ). We found that UK responders reported more infections and symptoms, a stronger financial hardship, and a stronger impact on health and the financial situation of family members. We found that responders of both countries reported an increase in psychological symptoms, especially depressive symptoms and anxieties. The global severity index (GSI) of the SCL was higher in UK responders compared to German responders. An alarming finding was that the percentage of people above clinical cut-off on the SCL-27 compared to a norm population had more than doubled for depressive, dysthymic and agoraphobic symptoms as well as for social phobias, and this increase was stronger in UK responders compared to German responders. We did not find differences in the SPQ or its subscales between the two countries. However, responders reported an increase of symptoms in about 9% with half of those reporting symptoms for the first time. Interestingly, despite the differences, UK responders were still more hopeful for a sooner end of the pandemic in their region, were less concerned about overall life stability and reported more positive changes due to the pandemic (e.g., time with the family, no commute, time for one-self).

In simple associative prediction models, we furthermore identified risk factors for the psychological impact of the pandemic. Being UK resident, female, younger, having a lower education, a worse pre-pandemic mental or physical health, as well as being more concerned about life stability, spending less time outside and reporting a stronger negative impact of the pandemic on the qualities of social contacts predicted higher scores of the GSI, as well as depressive, dysthymic symptoms as well as symptoms of anxiety. Higher scores on the SPQ total-score and its subdimensions were predicted by younger age, lower education, more substance (tobacco, vaping, marihuana) and media (social and video games), less sleep, less time spent outside, worse quality of social contacts, and a worse pre-pandemic mental and physical health.

To the best of our knowledge this is the first study showing a direct comparison of the psychological impact including schizotypy of the pandemic between two countries. There are several large scale studies reporting assessment of levels of depression, anxiety and stress related to COVID-19 comparing multiple countries and regions [15, 24]. This study identified prevalence and risk factors globally, but does not draw direct comparisons between different WHO-regions. Interestingly, however, Plomecka and colleagues (2020) report similar overall risk factors, such as being female, younger, less optimistic, and having worse social relationships and pre-pandemic.

A special focus needs to be drawn on developing and low- or middle-income countries, as those countries historically not only use a fraction of the global resources for mental health care and prevention [25], but also face a much harder impact of the economic consequences of the pandemic [26]; both of these aspects tremendously affect population mental health. In many low- and middle income countries, implementing restrictive measures in order to prevent the spread of the virus has a direct effect on the income of many day laborers, leaving them in direct fear of hunger for themselves and their families [27]. In high-income countries, increased mental health risks are, among other factors, linked to low socioeconomic status, low education, and over-crowed housing [28]. These aspects are highly prominent in low- and middle income countries which might further increase the risk for mental health problems. A recent review on the impact of the COVID-19 pandemic on mental health in low- and middle income countries across Asia and Africa [29] points out that most studies investigating this topic report increased levels of depression, post-traumatic stress disorder, adjustment disorders, addiction problems, sleep disorders, and anxiety disorders; the lack of thorough investigation of mental health in general and the poor quality of infrastructure for prevention and intervention remain pressing problems in low- and middle income countries.

Several studies investigate the psychological impact of the pandemic with a national focus. Two longitudinal studies conducted in UK populations [30, 31] show a general deterioration of mental health in April compared to before the pandemic. Both studies identify similar risk factors such as being female, younger of age and having pre-pandemic mental health conditions. The same is true for research conducted on German populations. A study by Bäuerle and colleagues [32] reports an increase in anxiety, depression and psychological distress with females and younger adults reported a stronger impact. Interestingly, Benke and colleagues [33] report similar effects but dissociate them from the governmental measures taken to control the pandemic. Another recent study [34] compared two countries, Poland and China, that differently enforced mask wearing during the initial stages of the pandemic and compared mental and physical health outcomes. For Poland, the country which less enforced mask wearing, the authors report higher levels of anxiety, depression and stress, as well as physical symptoms related to a COVID-19-infection.

Our study does not contain true pre-pandemic data. However, we assessed subjective measures of change questions on life circumstance and mental health question including the psychological questionnaires, asking participants to either report on that particular question three month ago or report whether symptoms had increased, decrease or stayed the same. In the UK population, we found a tripling of the percentage of people lying above cut-off compared to a norm population for depressive, dysthymic and agoraphobic symptoms, and a doubling on symptoms of social phobia and symptoms of mistrust. Similarly, Kwong and colleagues (2020) report a doubling of symptoms of anxiety in a UK sample. In the German responders, we found a doubling for depressive and dysthymic symptoms, for symptoms of social phobia and symptoms of mistrust. The increase in our study compared to Kwong et al (2020) might be due to the fact that the SCL-27 aims at high sensitivity, but low specificity on the individual symptoms. However, the increase is alarming, and requires actions for interventions.

Overall, our results match those of countries in a global comparison [15], and individual countries such as China [13, 35], Bangladesh [36], Brazil [37], South-Africa [38], Lebanon [39], Greece [40], Iran [41], Japan [42], India [43, 44], Italy [45] or Spain [46, 47]. Here, we provide a unique comparison of two economically and culturally similar countries. However, the governments of both countries followed different strategies in responding to the pandemic, whereas the German government implemented a prompt lock-down [16], the British government first discussed herd-immunity [48], causing a significant delay to implement the lock-down, which according to different predictive models has significantly increased the number of death in the UK [16]. At the time when we started the data collection the rise in cases in Germany was slowing down, whereas the cases in the UK were still increasing quickly, which may have influenced the results. The convenience sample nature of the participants is a limitation as it could also contribute to the observed results. Although our study does include participants with pre-existing mental illness (overall: 14.22%; UK:20.29%; Germany: 11.59%), it was not designed to specifically address mental health impact of the pandemic on those with severe and enduring mental illness, as this would require a more targeted study design. Although, the comparison of the two countries is still difficult, as both countries vary on a large number of factors not accounted for in this study that might have additionally contributed to the difference, it is likely that the burden of higher death rates and hospitalisations has increased the impact on mental well-being described in this study.

Interestingly, we find this dichotomy between a stronger financial and health impact of the pandemic on UK residents compared to German residents, and still a more optimistic judgment of the overall situation of the UK compared to the German residents. Further research would be needed to further investigate how pre-existing cultural attitudes contribute to these differences. We speculate there could be cultural differences in how likely people are to complain about their personal situation in a questionnaire, also there could be some linguistic difference in how these questions are understood by participants of the two countries. Another line of future enquiry could examine the role of such attitudes as the stereotypes of the British ‘Keep calm and carry on’ way of life [49] compared to the German stereotype of criticism and pessimism [50].

The ultimate question raised by our findings, and those of the many other studies investigating this important topic, is how to establish early interventions for mental health problems during a public health emergency? A number of reviews and opinion articles address this question in detail [4, 51–54]. The most important aspects proposed to date are psychoeducation and support for health care works, the detection of psychological problems or crises in the general population through online surveys and questionnaires, increased access to online consultations with health care professionals, as well as the development of intervention apps and online tools targeted for specific disorders such as anxiety disorder or depression.

### Limitations

This study has potential limitations. First, we used a purely online data collection methods, therefore, people without or with limited access to computers, or less well-versed using these methods would be excluded from the sample. However, in order maximally ease the accessibility of the questionnaire we provided an online version with smart-phone compatible formatting. Second, we used a snowball sampling method, therefore, the sample is not fully representable of the general population. The results of the study should therefore be interpreted considering the sample’s demographics. Third, comparing two countries is problematic as the countries vary on a large number of factors that are not and cannot be accounted for in detail. Therefore, any differences between the countries presented in this study might be linked to baseline differences. However, by specifically asking for a subjective change considering a pre-verses during-pandemic time-point, we minimised this confound. Fourth, we used a self-reporting survey without clinical in-person verifications. Social distancing measures complicate such verification. However, by using a completely voluntary and anonymous format, as well as standardised questionnaires we are minimising potential effects. And fifth, we are presenting simple logistic prediction models without testing for confounds and interactions. Although this approach may not present conclusive results, it does allow for comparison with other studies following the same approach, and to generate hypothesis for future research rather than definitive inference.

## Data Availability

The data used in this paper is available on personal request.

## Declarations

### Ethics approval and consent to participate

Ethical approval was obtained from the Ethical Commission Board of the Technical University Munich (250/20 S). All respondents included in the analyses provided informed consent. Our research was performed in accordance with the Declaration of Helsinki.

### Consent for publication

Not applicable.

### Availability of data and materials

The dataset generated during the current study are not publicly available while data collection is ongoing but are available from the corresponding author on reasonable request. The anticipated end of the data collection is December 2021.

## Authors’ Contributions

FK: administration of the project, conceptualisation, methodology, formal analysis and writing (original draft, review and editing), administration of the project. LR: formal analysis, editing of manuscript. GKM: methodology, and writing (review and editing).

## Acknowledgement

We thank the participants of the study who contributed their time and further circulated the survey. We thank Lorenz Mihatsch for the fruitful discussions and Sarah Daimer for help with the revision of the manuscript

## Supplementary materials

**Suppl. Figure 1.**
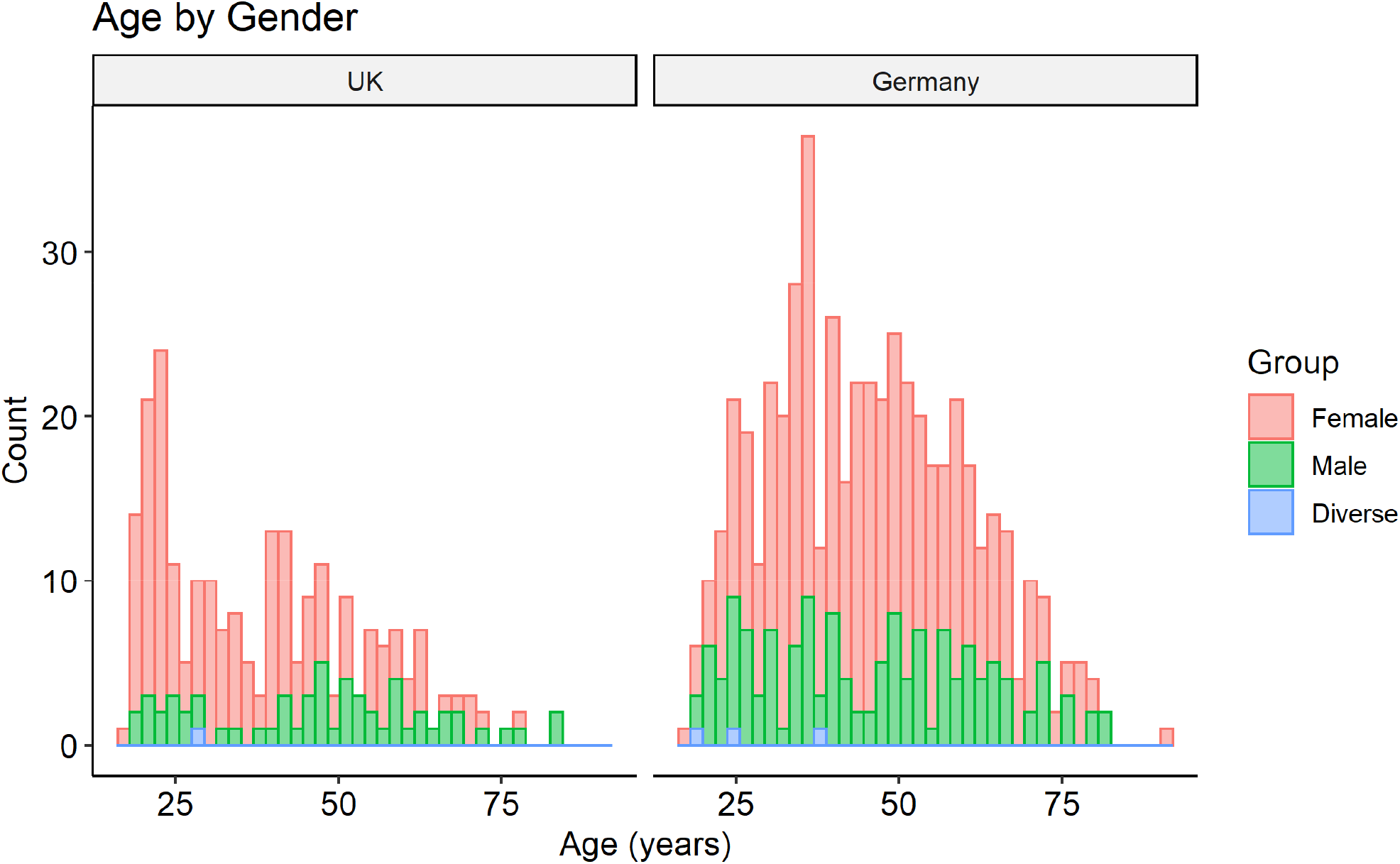
Distribution of age by gender for the UK and Germany separately.

**Supp. Figure 2.**
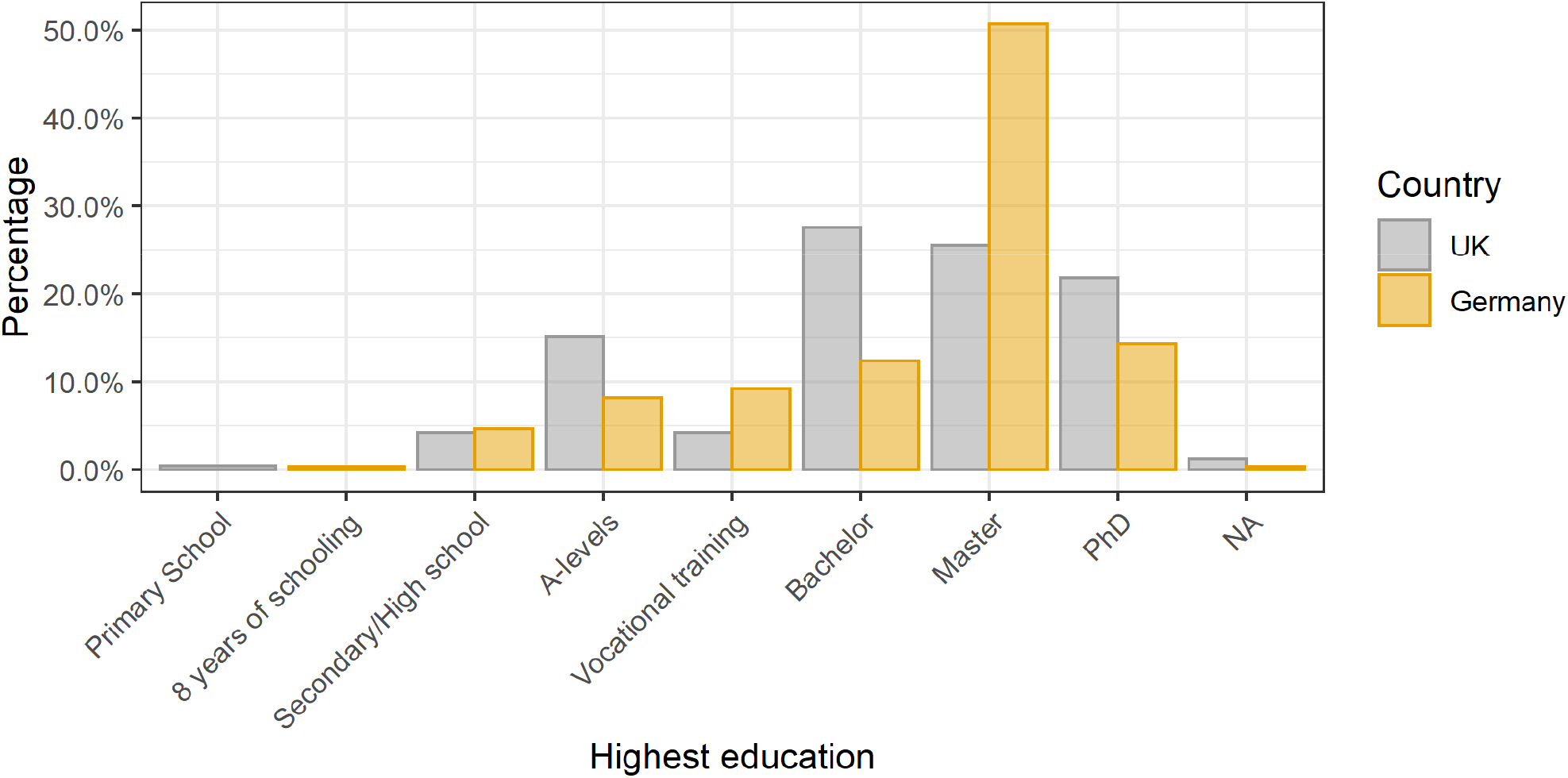
Distribution of highest personal education by country.

**Supp. Figure 3.**
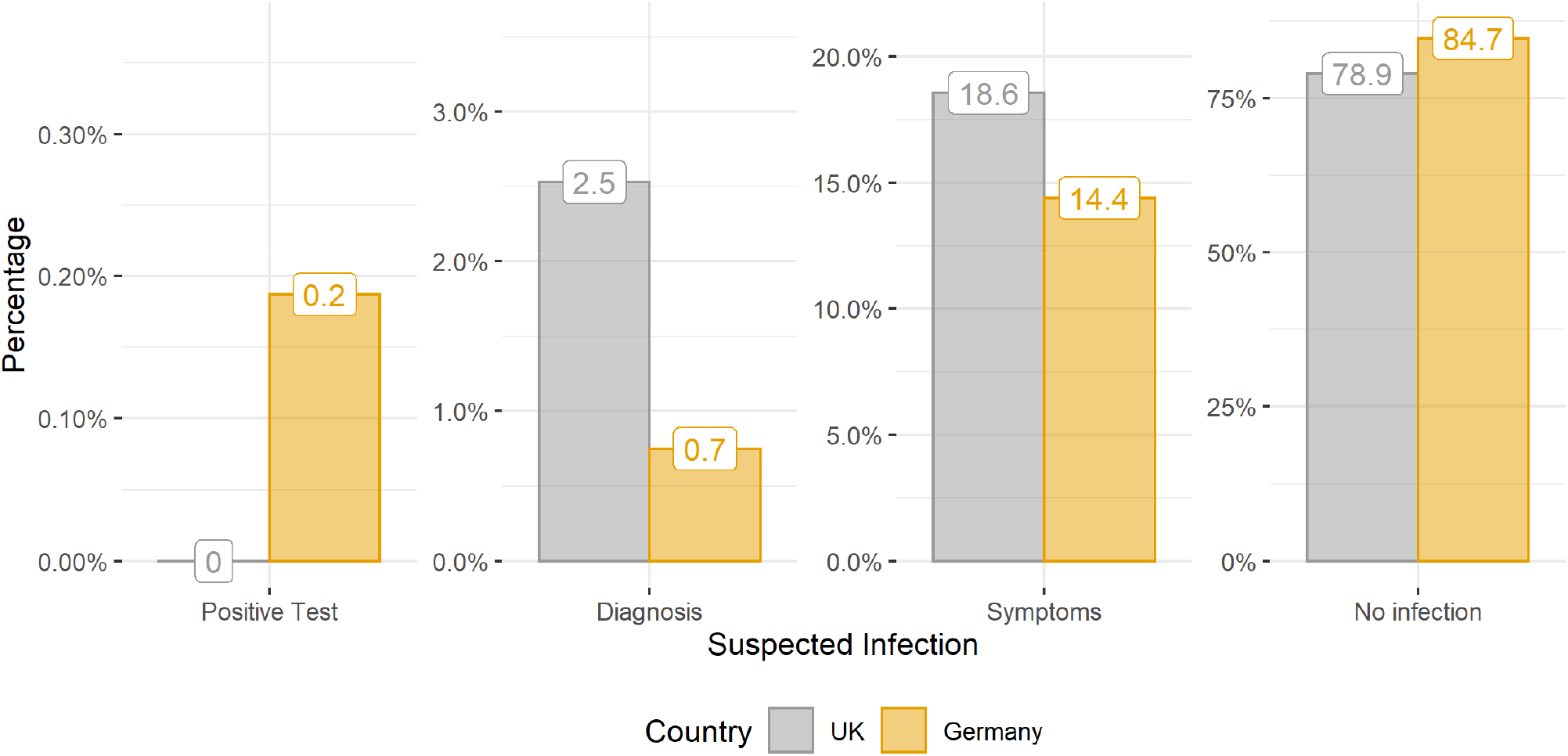
Percentage of suspected infection by country, presented in varying scales.

**Supp. Figure 4.**
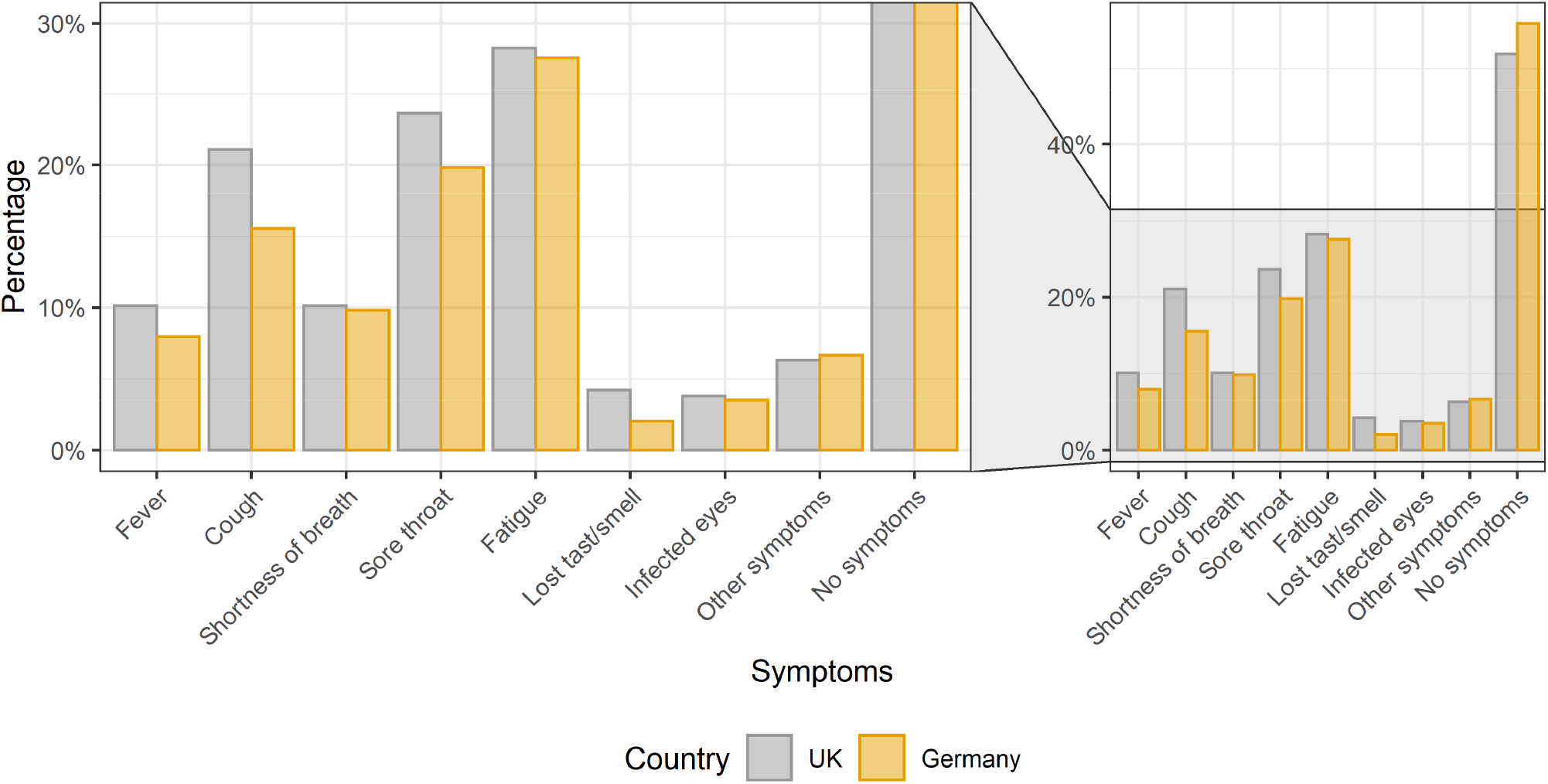
Distribution of symptoms by country, with close-up presented in the left panel.

**Supp. Figure 5.**
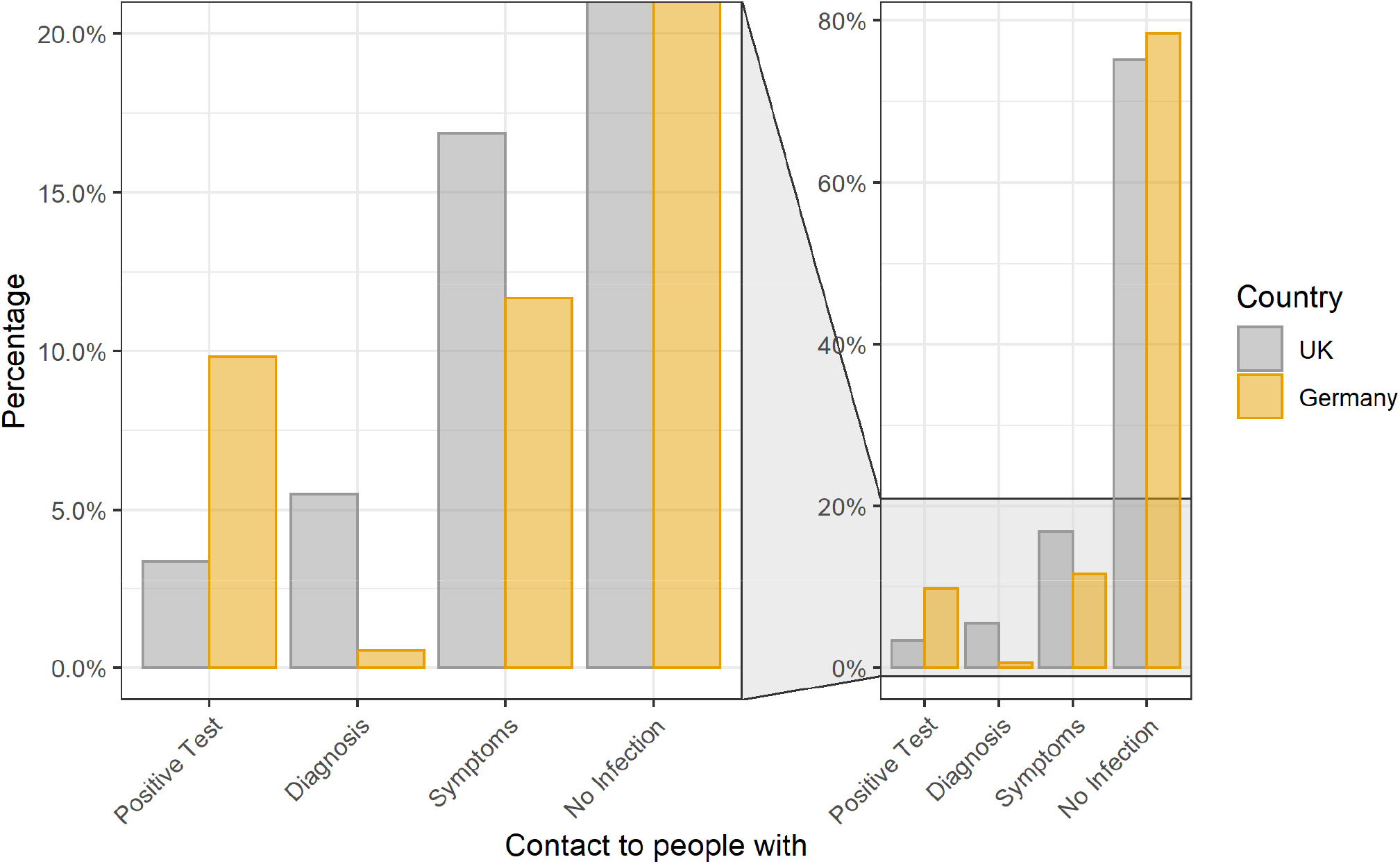
Distribution of contacts to people with possible infections by country, with close-up presented in the left panel.

**Supp. Figure 6.**
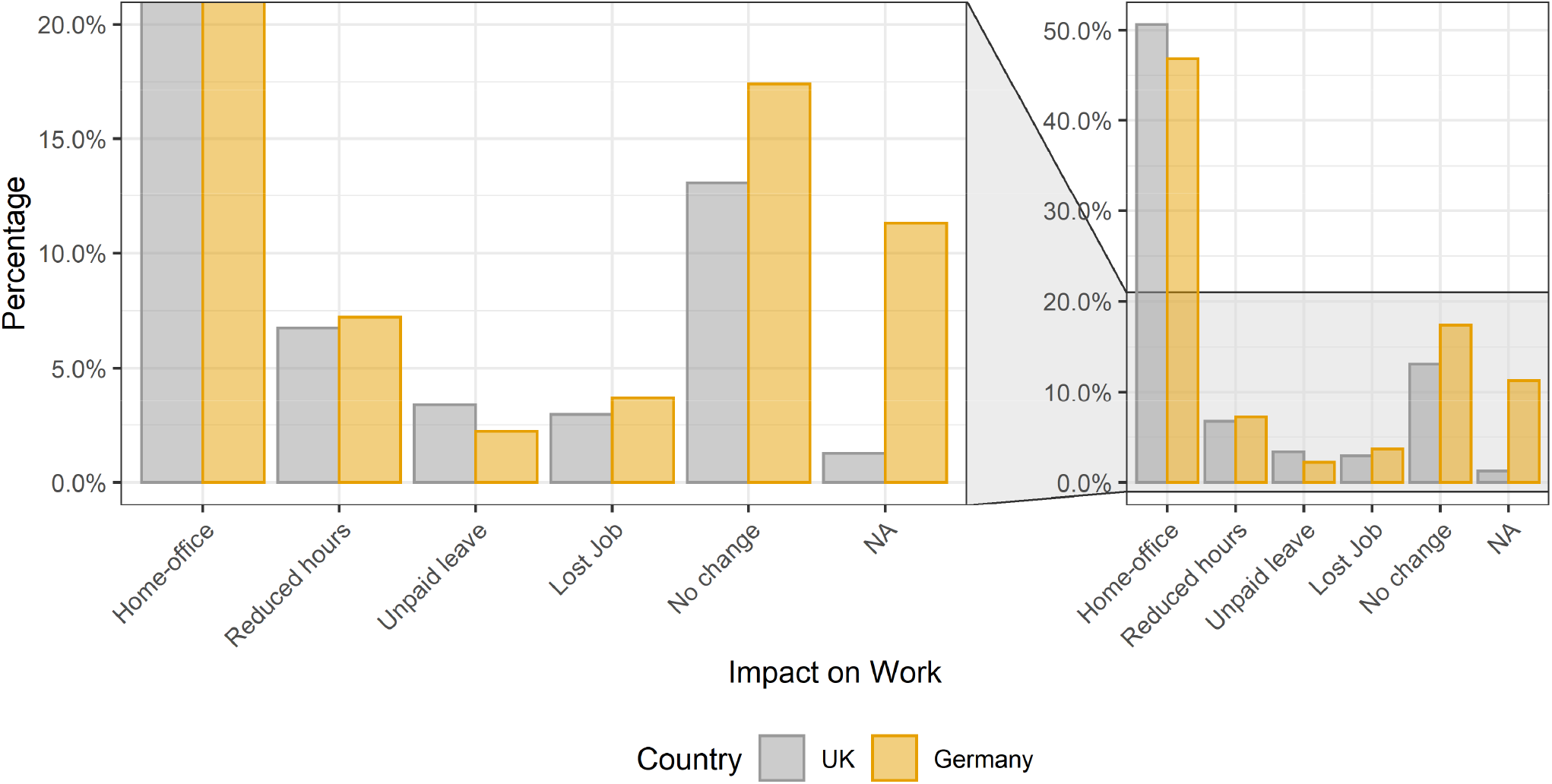
Impact on working situation by country, with close-up presented in the left panel.

**Supp. Figure 7.**
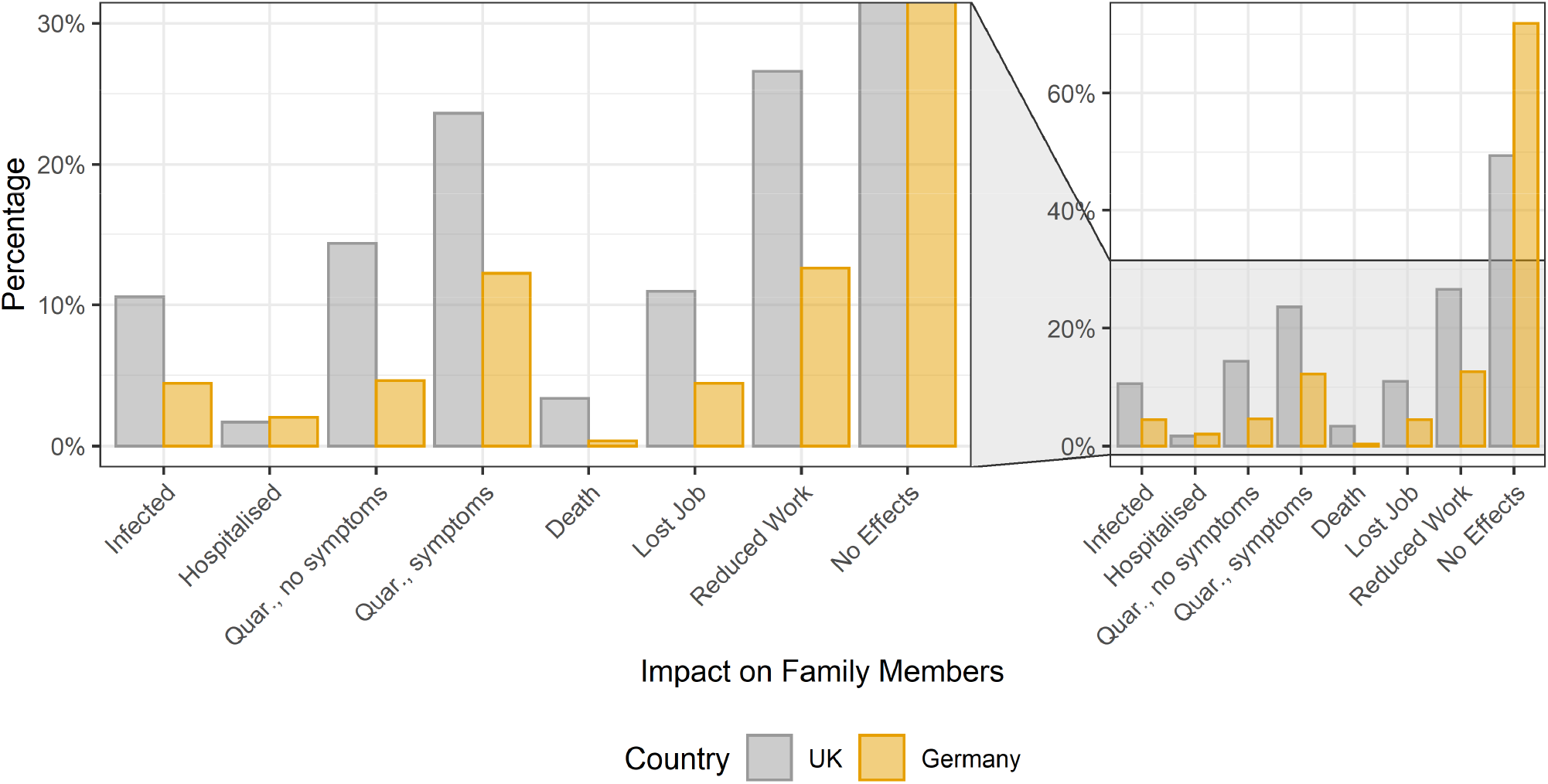
Impact on family by country, with close-up presented in the left panel.

**Supp. Figure 8.**
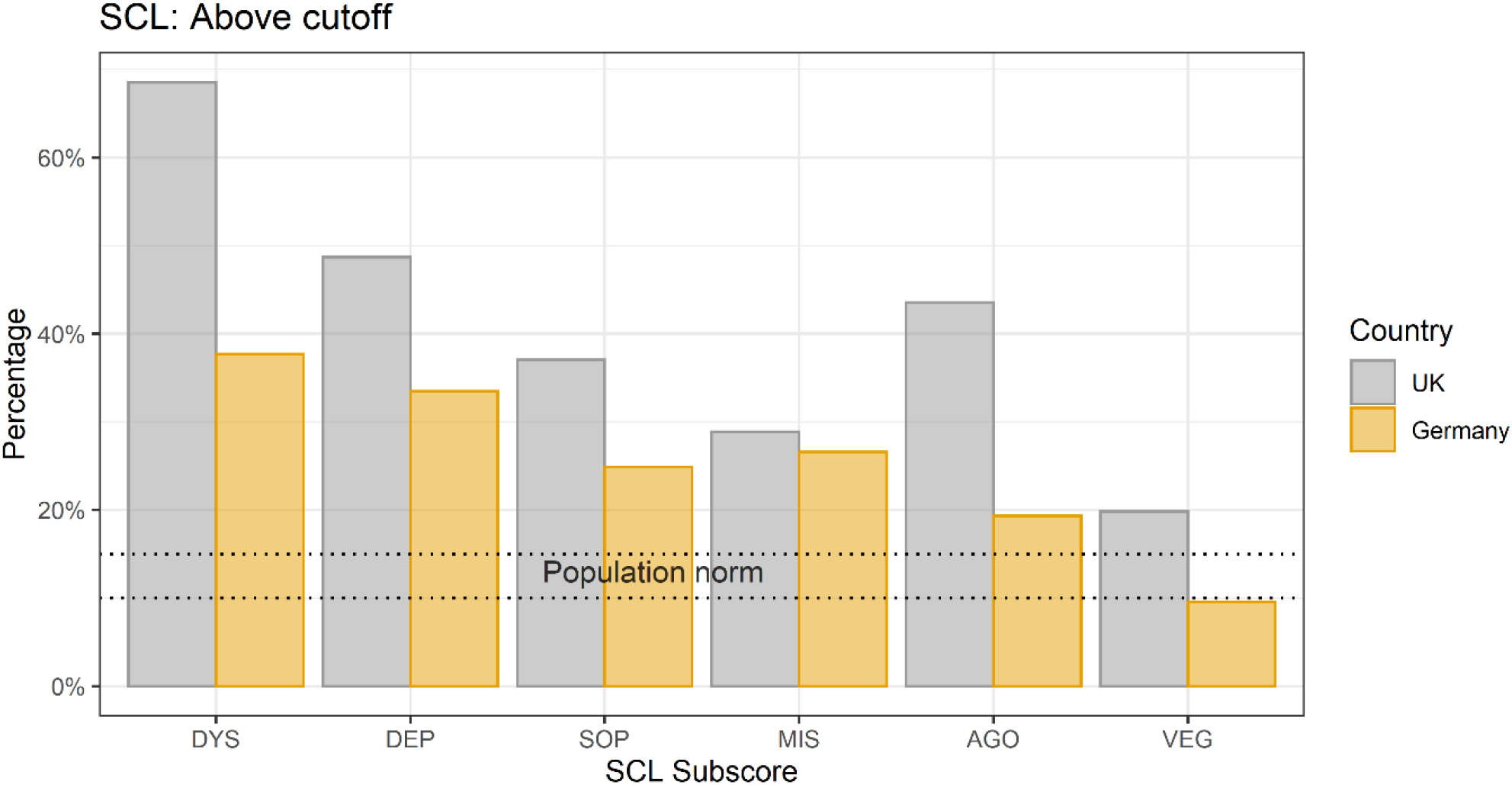
Percentage of responders above clinical cut-off separately for UK and Germany. Dotted lines represent the percentage of the norm population above threshold. DYS: dysthymic symptoms, DEP: depressive symptoms, SOP: symptoms of social phobia, MIS: symptoms of mistrust, AGO: agoraphobic symptoms, VEG: vegetative symptoms.

**Supp. Figure 9.**
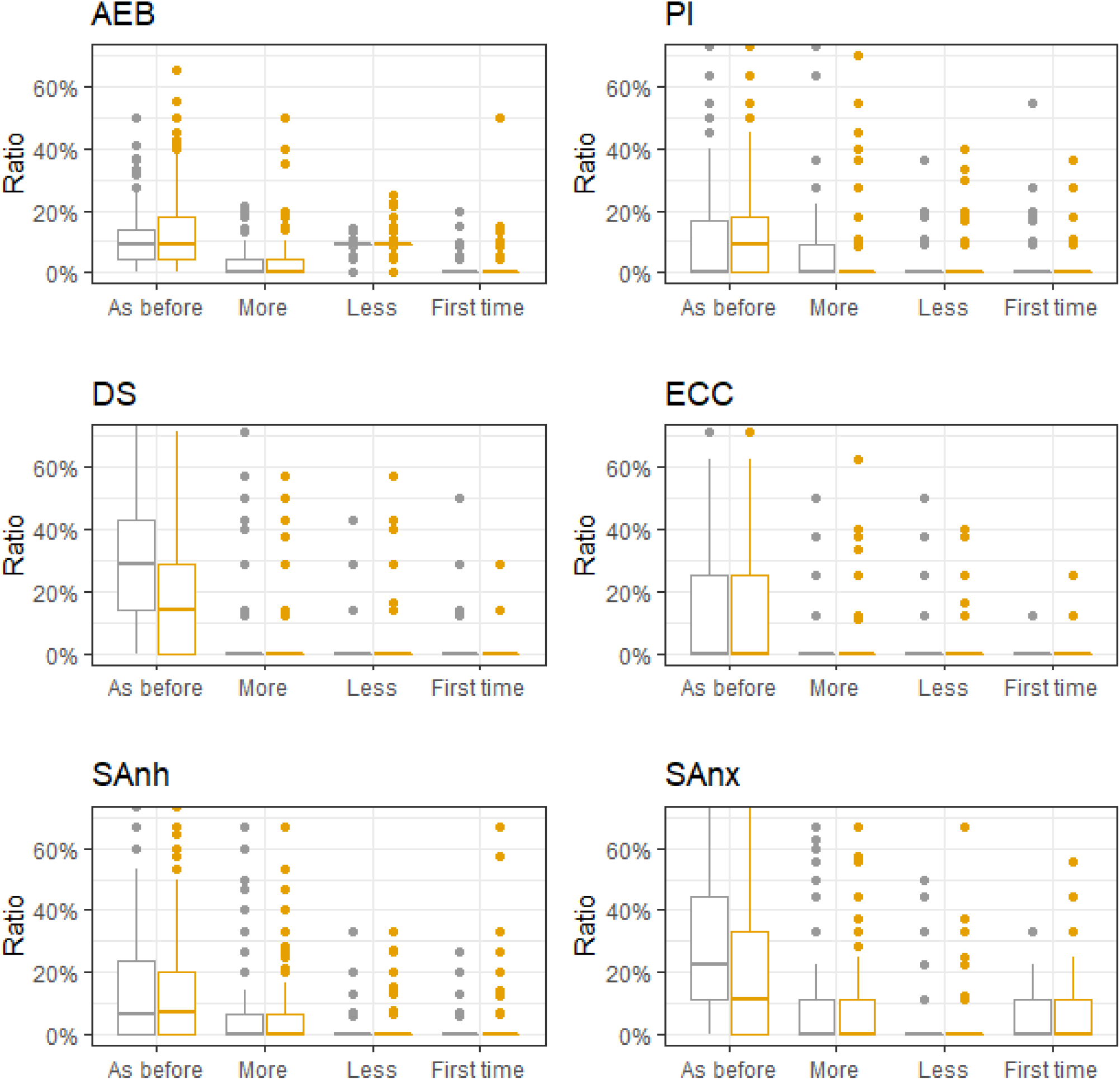
Each boxplot shows the subjective change of one of the sub-dimensions (experiences and beliefs (AEB), social Anhedonia (SAnh), paranoid ideation (PI), social anxiety (SAnx), eccentricity (Ecc), and disorganised speech (DS)) of the SPQ during the pandemic, separately for Germany (yellow-orange) and the UK (grey).

**Supp. Figure 10.**
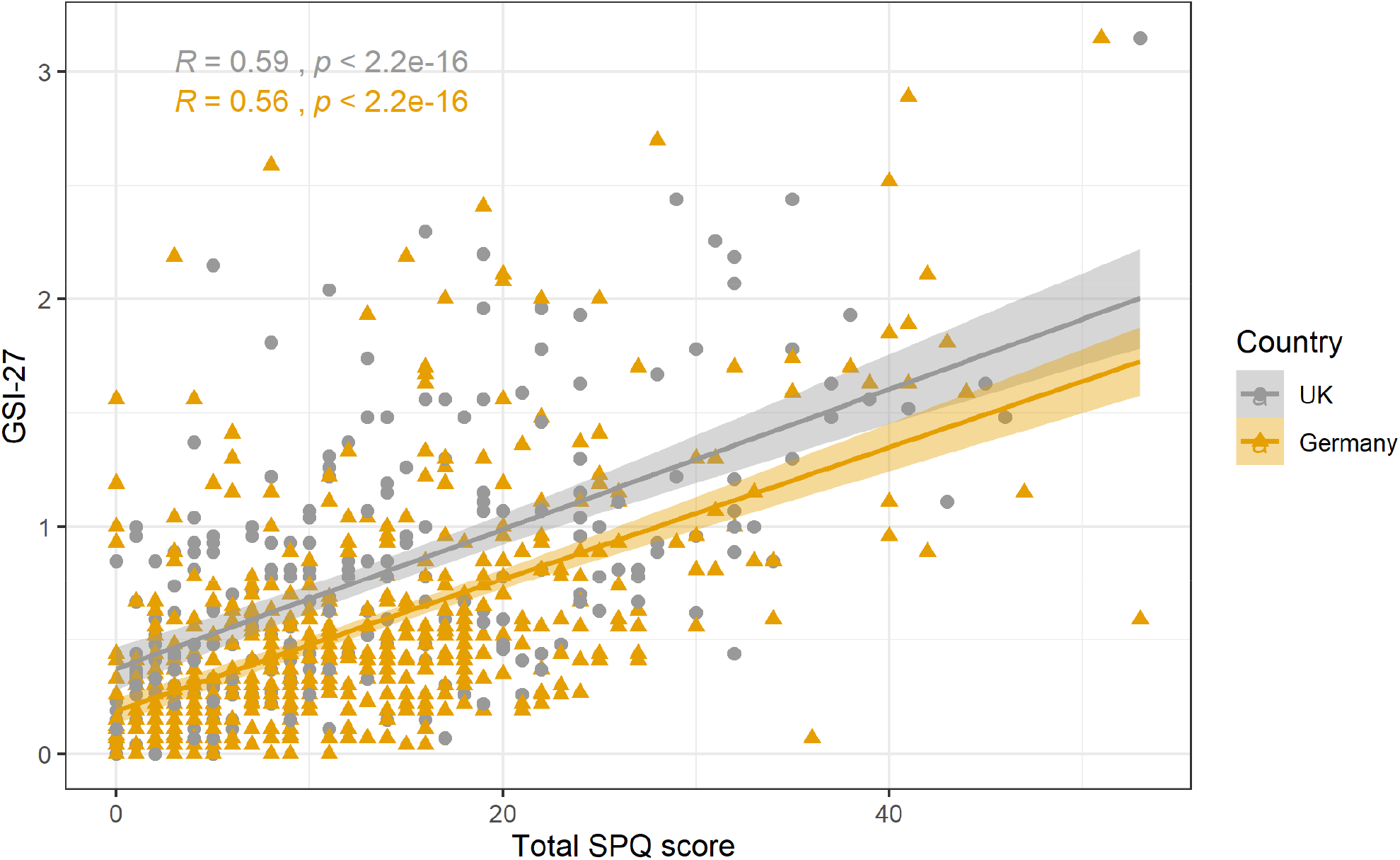
Pearson correlation between GSI-27 and total SPQ score, separately per country. GSI-27 and SPQ total reveal a strong positive correlation.

